# Optic nerve tortuosity and displacements during horizontal eye movements in healthy and highly myopic subjects

**DOI:** 10.1101/2020.10.07.20208397

**Authors:** Xiaofei Wang, Stanley Chang, Jack Grinband, Lawrence A. Yannuzzi, K. Bailey Freund, Quan V. Hoang, Michaël J. A. Girard

## Abstract

**Purpose:** (1) To assess the morphology and 3D displacements of the eye globe and optic nerve (ON) in adduction/abduction using magnetic resonance imaging (MRI). (2) To assess differences between healthy emmetropic and highly myopic (HM) subjects.

**Methods:** MRI volumes of both eyes from 18 controls and 20 HM subjects in primary gaze, abduction and adduction (15°) were postprocessed. All ONs were manually-segmented and fitted to a 3D curve to assess ON tortuosity. ON displacements were evaluated in 4 quasicoronal planes which were perpendicular to the ON in primary gaze and 3 mm apart.

**Results:** Axial length was higher in the HM group (28.62±2.60 vs 22.84±0.89 mm; p<0.0001). Adjusted ON tortuosities (i.e., ON tortuosities estimated before myopia onset) were lower in HM eyes (0.9063±0.0591) versus controls (1.0152±0.02981) in primary gaze, adduction (0.9023±0.05538 versus 1.0137±0.0299) and abduction (0.9100±0.0594 versus 1.0182±0.0316); p<0.0001 for all cases. In all eyes, ON displacements in adduction were significantly different from those in abduction in the naso-temporal direction (p<0.0001 in all planes) but not in the supero-inferior direction. ON displacements in the posterior segments of the ON were smaller in the HM group in both gaze directions and were larger in the anterior-most ON segment in adduction only.

**Conclusions:** The adjusted tortuosity of the ON was significantly lower in HM eyes, suggesting that eyes destined toward HM exhibited higher ON traction forces during eye movements before the onset of myopia. Our ON metrics may be valuable to explore a potential link between eye movements and axial elongation.

## INTRODUCTION

Myopia is the most common refractive error in young population and is characterized by an axial elongation of the eye over time. Currently, there is a high prevalence of myopia and high myopia (HM) worldwide, especially in East and Southeast Asia.^1,2^ HM is highly correlated with pathological myopia, which could lead to complications such as myopic macular degeneration, choroidal neovascularization and retinal detachment.^3^

Myopia has a multifactorial aetiology. Both genetic and environmental factors may contribute to myopia onset and progression.^4^ The recent global rise of myopia prevalence indicates that environmental factors could be critical in the development of myopia. The major environmental risk factor for myopia is education pressure, which includes long duration of near work and limited time outdoors during daylight hours.^5,6^ A number of studies have shown that the link between near work and myopia is consistent.^7–9^ Although the association between myopia and near work has long been established, its mechanism is still unclear.

During near work, accommodation and convergence (combined with other types of eye movements such as downward gaze) of the eye occur simultaneously. Studies have shown that accommodation could induce a small transient increase in axial length.^10–13^ However, after removing the effect of accommodation, a few studies have found that eye movement alone during near work (such as convergence and/or downward gaze) could result in significant axial elongation.^14–16^ These studies have speculated that forces generated by the extraocular muscles during eye movements may be the cause of axial elongation in convergence. However, the directions of extraocular muscle forces are not along the axial direction, especially in light of the muscle pulley theory,^17^ thus it is unlikely that these forces would cause scleral remodelling to yield posterior expansion of the globe. Our previous studies have shown that the optic nerve (ON) traction force exerted on the eye globe during eye movements could deform the optic nerve head (ONH), and such effects were more pronounced in adduction than in abduction.^18–20^ Moreover, the ON traction force is on the same order of magnitude as the extraocular muscle forces and acting along the axial direction.^21^ Based on the above observations, we hypothesized that the ON traction force could play a role in axial elongation in myopia.^21^

The ON traction force during eye movements is determined by many factors, including the stiffness, morphology, microstructural collagenous organization,^22^ and spatial arrangements of ocular tissues.^21^ Although it is not yet feasible to measure the ON traction force *in vivo*, the morphology and the displacements of the eye globe and ON during eye movements can be assessed with magnetic resonance imaging (MRI). These factors may reflect the magnitude and the effects of ON traction. While our group and others have studied ON morphology in glaucoma eyes^20,23^, no studies have yet investigated ON morphology and displacements in HM subjects.

The aim of this study was to compare the differences in the morphology and displacement patterns of the globe and ON during eye movements in normal and HM eyes.

## METHODS

### Subject Recruitment & Clinical Tests

Twenty HM subjects and 18 healthy emmetropic controls were included in this study. All HM subjects had at least one eye with an axial length of > 27 mm on partial coherence interferometry (Carl Zeiss Meditec, Inc, Dublin, CA) and a clinical diagnosis of staphyloma. HM subjects underwent a complete clinical examination including dilated fundus examination, B-scan ultrasonography (Quantel Aviso, Rockwall, TX) and swept source optical coherence tomography (SS-OCT, DRI OCT-1, Atlantis, Topcon, Oakland, NJ). All subjects had a best-corrected visual acuity of 20/40 or greater in at least one eye.

Staphylomas were classified into four types by a single grader (QVH): macular staphyloma (Type-1), posterior pole staphyloma (Type-2), compound staphyloma (Type-3; two bulges) and peripapillary staphyloma (Type-4).

This study complied with the Health Insurance Portability and Accountability Act of 1996 and adhered to the tenets of the Declaration of Helsinki. Institutional Review Board (IRB)/Ethnics Committee approval was obtained from Columbia University. All patients signed a written consent form before initiation of the study-specific procedures.

### Magnetic Resonance Imaging and Processing

Each subject’ s orbital regions were imaged with a Phillips 3T Achieva scanner with an 8-channel phased-array head coil using a fat-suppressed, axial T2-weighted volumetric scanning sequence (1024 x 1024 matrix, number of slices = 50, pixel size = 0.215 mm, slice thickness = 1.0 mm, TR = 4000 ms, TE = 475 ms). **Figure 1A** and **1B** show representative axial MRI images obtained in this study.

**Figure 1.**
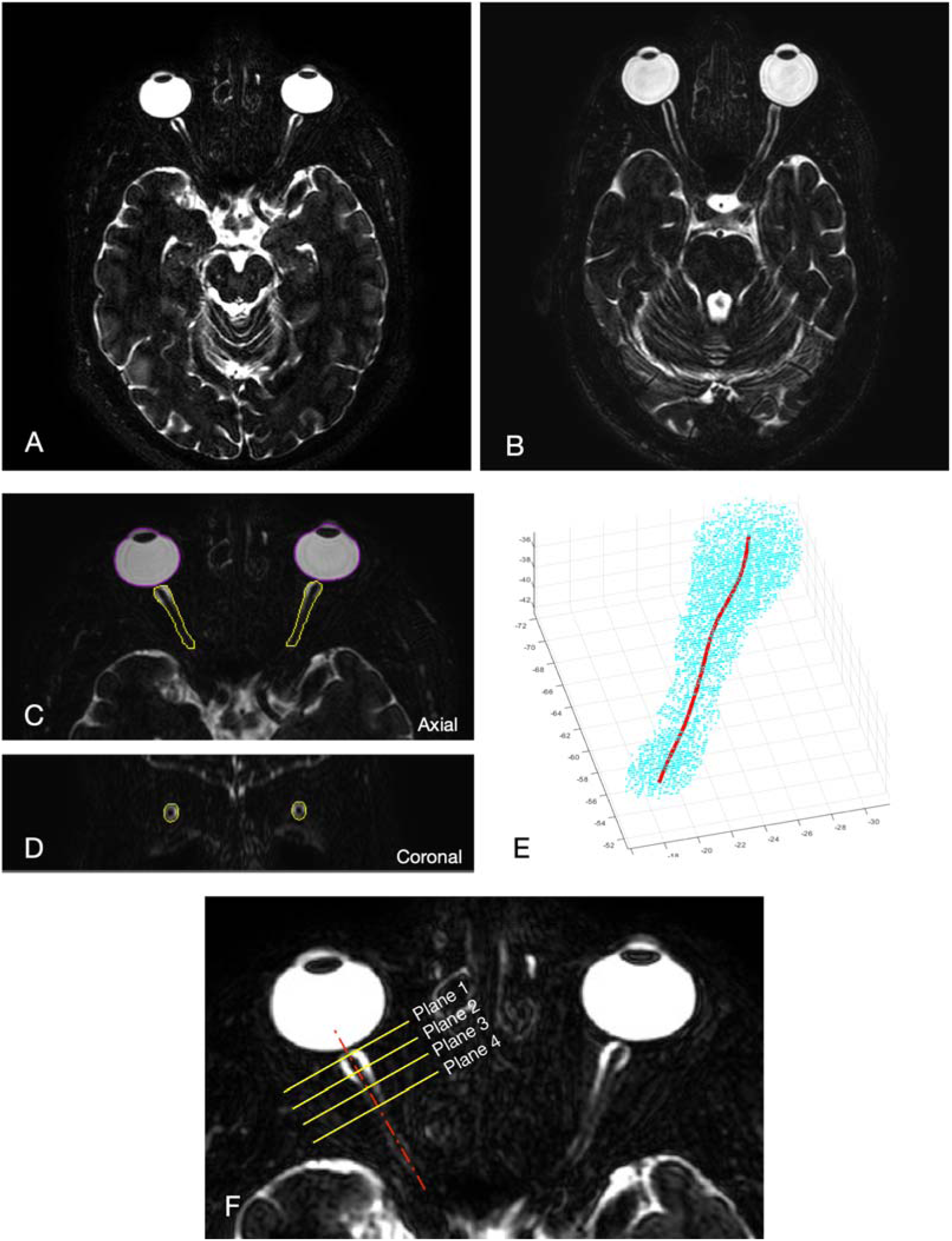
Examples of axial MRI images of an emmetropic control subject **(A)** and a highly myopic subject **(B)** obtained in this study. Globes and optic nerves (ON) were then manually segmented, as shown in axial (**C**) and coronal (**D**) views. All three MRI volumes (primary, left and right gaze) were co-localized before segmentation. (**E**) Segmented ONs were fitted to a 3D spline curve. **(F)** Illustration of the 4 quasicoronal planes where ON displacements were measured. These 4 planes were perpendicular to the ON central line in primary gaze and 3 mm apart.

During the MRI procedure, the subjects were instructed to direct their gaze on one of the three visual targets viewable via a standard head-coil-based mirror, allowing image acquisition in the primary position (baseline), and on left and right gaze of 15°.

### MRI Volume Processing

MRI volumes of both eyes for each subject in primary gaze, abduction and adduction of 15° were analysed. All MRI volumes were resampled from 1024×1024×50 to 1024×1024×200 voxels to obtain a smoother representation of the ON boundaries. Left and right gaze MRI volumes were reoriented to align with the baseline volume to remove any possible head rotation and to allow comparison of globe and ON positions in the same coordinate system. The reorientation included rigid translation and rotation transformations of the MRI volume with Amira (version 6.0; FEI) using a voxel-based algorithm that maximized mutual information between two given volumes.^24^ In the reorientation process, the orbital regions were cropped out to remove the effects of eye rotations on the 3D volume registration, allowing a precise alignment of MRI volumes solely using the landmarks of the brain. Images were manually-segmented in coronal, axial and sagittal views to isolate both the globe and ON pixels (**Figures 1C,1D**).

### Measurements of Eye Globe Length and Displacements

The length of each globe was measured from 3D MRI scans automatically using delineated images, which was used as a surrogate for the axial length. In this study, MRI-based globe length measurements correlated well with axial length measurements obtained by IOLMaster.

Globe displacements were calculated based on the location of the globe center at each gaze positions. The globe center was defined as the geometric center of all segmented globe pixels.

### Measurements of ON Tortuosity

First, each ON was sliced by around 100 coronal slices (0.215 mm apart). For each coronal slice, we computed the geometric center from the ON contour points within that plane.

All obtained points (geometric centers) were then fitted to a 3D curve (spline) to form the ON central line (**Figure 1E**). The tortuosity of the ON was defined as the length of the ON central line segment (15 mm long from the globe-ON junction) divided by the distance between the two end-points. The adjusted ON tortuosity was defined as the ON tortuosity, but estimated prior to the onset of myopia. For instance, for a given HM eye, the adjusted ON tortuosity represents the tortuosity of the ON when the eye had a normal axial length at a prior point in time before the onset of myopia. It was defined as the length of the ON central line segment (15 mm) divided by a hypothetical distance of this ON segment when the axial length was normal (22.84 mm, the average value of all normal eyes). Conservative assumptions were made to ensure we did not under-estimate such adjusted tortuosity values. See supplementary material A for further details.

### Measurements of ON Displacement

ON displacements were evaluated in 4 quasicoronal planes (spaced in 3 mm intervals), which were oriented perpendicular to the ON central line in primary gaze (**Figure 1F**). Specifically, on each plane, the shift of the ON central line at each gaze position was calculated along the naso-temporal (X) and supero-inferior (Z) axes.

### Measurements of ON Insertion Angle

The ON insertion angle was defined as the angle between the limbal plane and the ON insertion plane, which should correlate to the clinical observation of optic nerve disc tilt. The limbal plane was defined by a 2D plane that was fit through the limbus and center of the lens in MRI volumetric renderings using an automatic algorithm (**Figure 2A**).^25^ The ON insertion plane was defined by fitting a plane to peripapillary scleral points within a 4-mm in diameter region centered on the ON-globe junction (**Figure 2B**).

**Figure 2.**
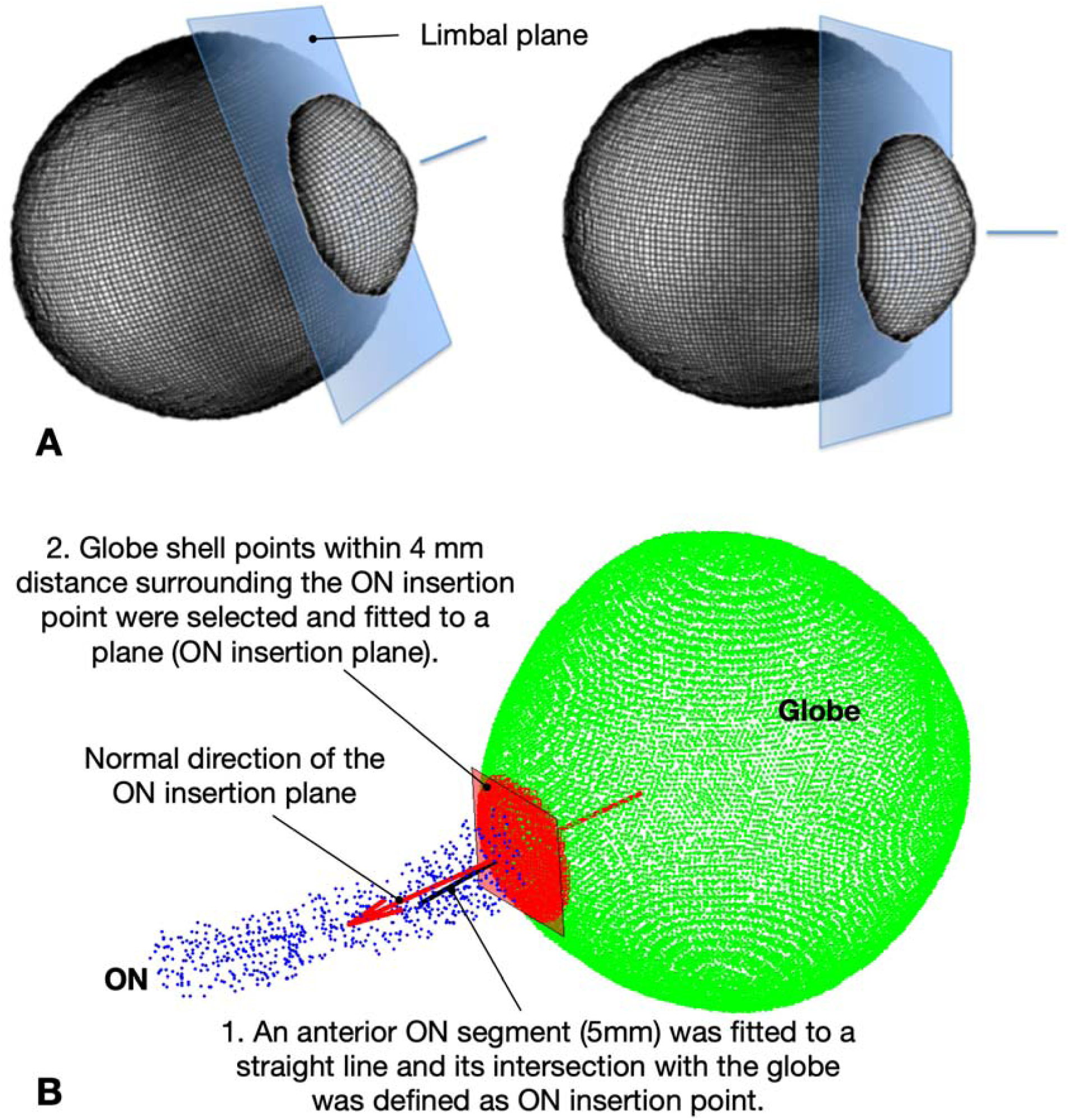
**(A)** The optic nerve (ON) insertion angle was defined as the angle between the normal from the limbal plane (blue) and that from the ON insertion plane (red). The limbal plane was defined by a 2D plane that was fit through the limbus and center of the lens from MRI volumetric renderings using an automatic algorithm. **(B)** The ON insertion plane was defined by fitting a plane to peripapillary scleral points contained within a 4-mm in diameter region centered on the ON-globe junction

### Statistics

All measured parameters were compared between control and HM eyes. As data from both eyes were used, a linear mixed model was employed to adjust for inter-eye correlations. Correlations between ON insertion angles and other parameters were also evaluated using linear mixed models. A Tukey’ s multiple comparison test was used to compare insertion angles among different staphyloma types in HM eyes. All statistical analyses were performed using R (R Foundation, Vienna, Austria). P < 0.05 was considered as statistically significant.

## RESULTS

### Subjects

For this study, 18 normal controls (10 female and 8 male) and 20 HM (12 female and 8 male) subjects were included. The age of normal and HM subjects was 26.57±2.35 and 55.80±15.49 years, respectively. The axial length of HM cases (28.62±2.60 mm) was significantly greater than that of normal controls (22.84±0.89 mm) (P<0.0001). For HM eyes, there were 9 Type-1, 17 Type-2, 6 Type-3, 1 Type-4 staphylomas and 7 with no staphyloma.

### ON Morphology

**Figure 3** shows the relative location of the averaged ON central lines within the sagittal plane of control and HM subjects in primary gaze, after referencing all eyes to the same ON-globe junction. ONs of HM subjects were located more inferiorly than those of emmetropic controls. Specifically, in the 3 coronal planes that are located 5, 10, and 15 mm posterior to the ON-globe junction, the supero-inferior location of ONs was statistically different between two groups (p < 0.0001).

**Figure 3.**
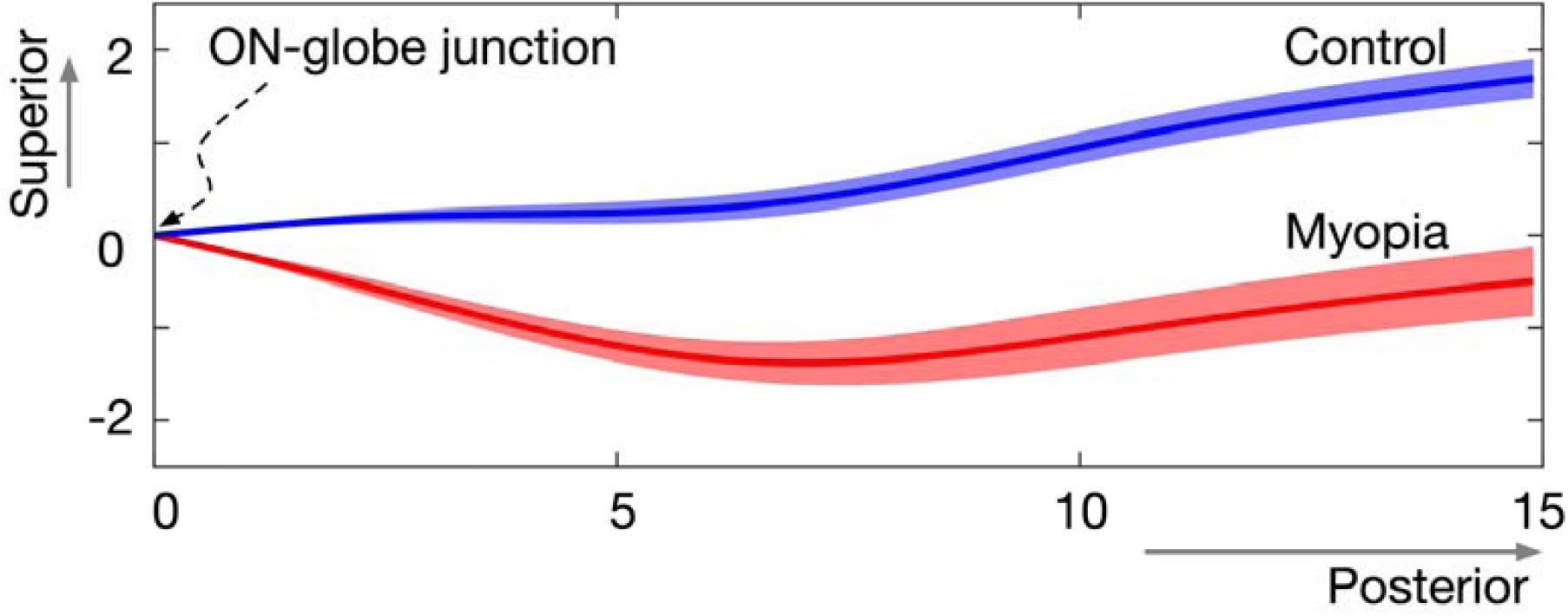
Average optic nerve location (mean ± standard error of the mean) in the sagittal plane for control and high myopia subjects. Unit: mm.

Raw (top panel) and adjusted (bottom panel) ON tortuosities in baseline, adduction, and abduction of 15° are summarized in **Figure 4** for both groups. In all gaze positions, the ON tortuosities of HM eyes were always significantly higher than those of the emmetropic controls (average value for all gaze positions: 1.0436 ± 0.0418 in HM eyes versus 1.0151 ± 0.01917 in controls). The adjusted ON tortuosities of HM eyes were significantly lower than those of the emmetropic controls in primary gaze (0.9063 ± 0.0591 versus 1.0152 ± 0.02981), adduction (0.9023 ± 0.05538 versus 1.0137 ± 0.0299) and abduction (0.9100 ± 0.0594 versus 1.0182 ± 0.0316); p<0.0001 for all cases.

**Figure 4.**
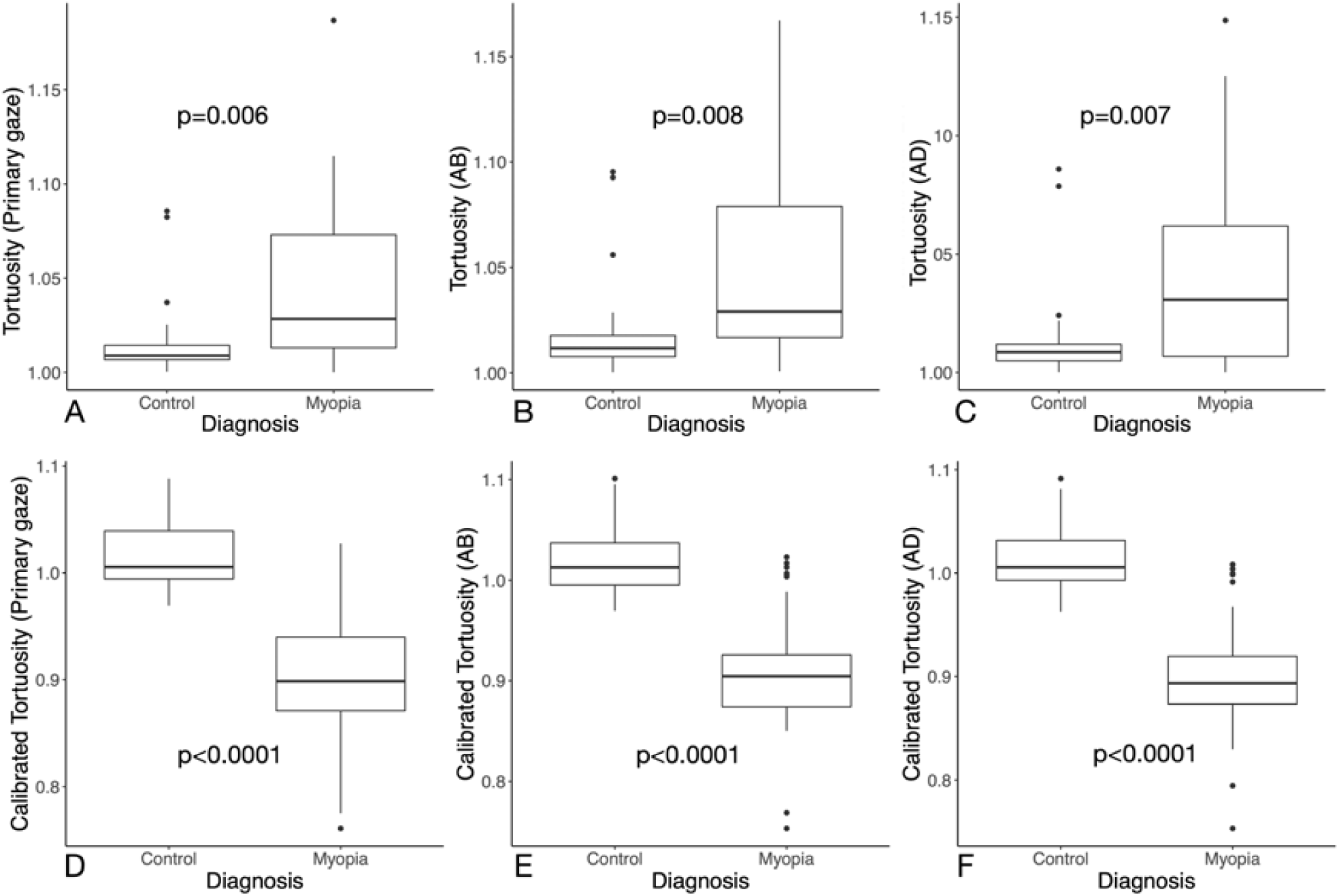
Optic nerve tortuosity in primary gaze **(A, D)**, abduction **(B, E)** and adduction **(C, F)** in control and high myopia groups. Top panel depicts observed measurements and bottom panel depicts the adjusted optic nerve tortuosity. AB: abduction, AD: adduction, ON: optic nerve.

### Globe Displacements

In the naso-temporal (X) direction (**Figure 5A)**, globe displacement was significantly higher for emmetropic eyes in both adduction and abduction. However, in the antero-posterior (Y) and supero-inferior (Z) directions, no statistical significant differences were found between the emmetropic controls and HM eyes **(Figures 5B, 5C**).

**Figure 5.**
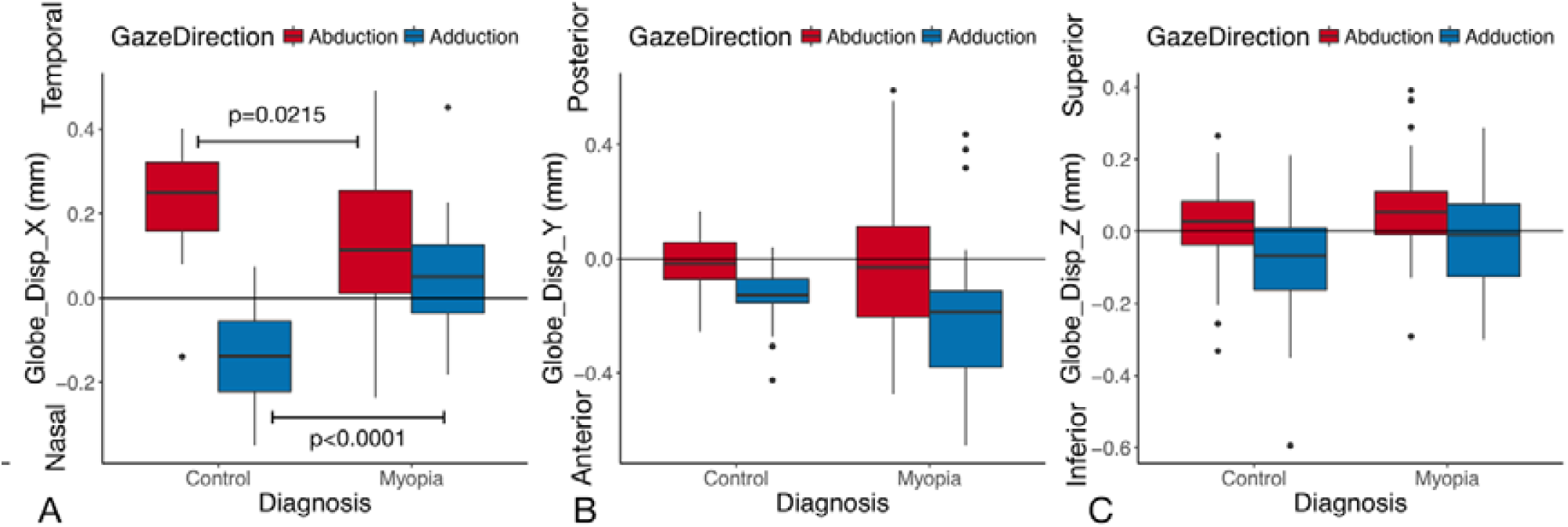
Globe displacements in control and myopia groups in **(A)** naso-temporal (X), **(B)** antero-posterior (Y) and **(C)** supero-inferior (Z) directions. Globe displacement in naso-temporal direction was significantly larger for control group in both adduction and abduction.

### ON Displacements

In all eyes, ON displacements in adduction were significantly different from those in abduction in the naso-temporal direction (p<0.0001 in all 4 planes) but not in the supero-inferior direction (p>0.26) (**Table 1**). ON moving distances (without considering directions) in the axial plane were similar in adduction and abduction (p>0.28 in all 4 planes). ON displacements in plane 3 and 4 were smaller in the HM group in both gaze directions and were larger in plane 1 in adduction only (**Table 2** and **Figure 6**).

**Table 1.** Comparisons of ON displacements in the naso-temporal and supero-inferior directions between adduction and abduction.

| Directions | Plane | Adduction | Abduction | p-value |
| --- | --- | --- | --- | --- |
| Temporal (+)<br>Nasal (-) | Plane 1 | 1.725<br>(0.386) | -1.720<br>(0.452) | **< 0.0001** |
|  | Plane 2 | 1.329<br>(0.353) | -1.347<br>(0.391) | **< 0.0001** |
|  | Plane 3 | 0.950<br>(0.391) | -0.905<br>(0.434) | **< 0.0001** |
|  | Plane 4 | 0.598<br>(0.363) | -0.566<br>(0.446) | **< 0.0001** |
| Superior (+)<br>Inferior (-) | Plane 1 | -0.006<br>(0.596) | -0.008<br>(0.633) | 0.2674 |
|  | Plane 2 | -0.004<br>(0.492) | -0.010<br>(0.413) | 0.6358 |
|  | Plane 3 | -0.026<br>(0.391) | -0.045<br>(0.397) | 0.7670 |
|  | Plane 4 | -0.041<br>(0.344) | -0.021<br>(0.359) | 0.4880 |

**Table 2.** Statistical results of ON displacements in 4 quasicoronal planes. Measurements were summarized as Mean (SD) except for p-values.

| Directions | Plane | Adduction |  |  | Abduction |  |  |
| --- | --- | --- | --- | --- | --- | --- | --- |
|  |  | Control | Myopia | p-value | Control | Myopia | p-value |
| Temporal (+)<br>Nasal (-) | Plane 1 | 1.580<br>(0.244) | 1.862<br>(0.445) | **0.0091** | -1.738<br>(0.253) | -1.703<br>(0.585) | 0.7519 |
|  | Plane 2 | 1.348<br>(0.253) | 1.311<br>(0.430) | 0.7349 | -1.422<br>(0.281) | -1.277<br>(0.466) | 0.2210 |
|  | Plane 3 | 1.075<br>(0.280) | 0.831<br>(0.444) | **0.0434** | -1.111<br>(0.369) | -0.711<br>(0.403) | **0.0022** |
|  | Plane 4 | 0.717<br>(0.285) | 0.486<br>(0.396) | **0.0295** | -0.785<br>(0.476) | -0.360<br>(0.299) | **0.0003** |
| Superior (+)<br>Inferior (-) | Plane 1 | -0.016<br>(0.339) | 0.003<br>(0.769) | 0.8076 | 0.010<br>(0.320) | -0.025<br>(0.832) | 0.8142 |
|  | Plane 2 | -0.104<br>(0.319) | 0.091<br>(0.601) | 0.1075 | -0.026<br>(0.333) | 0.005<br>(0.481) | 0.7690 |
|  | Plane 3 | -0.069<br>(0.314) | 0.015<br>(0.453) | 0.3811 | -0.041<br>(0.356) | -0.048<br>(0.438) | 0.9609 |
|  | Plane 4 | -0.051<br>(0.308) | -0.032<br>(0.379) | 0.8315 | -0.042<br>(0.365) | -0.002<br>(0.357) | 0.6748 |

**Figure 6.**
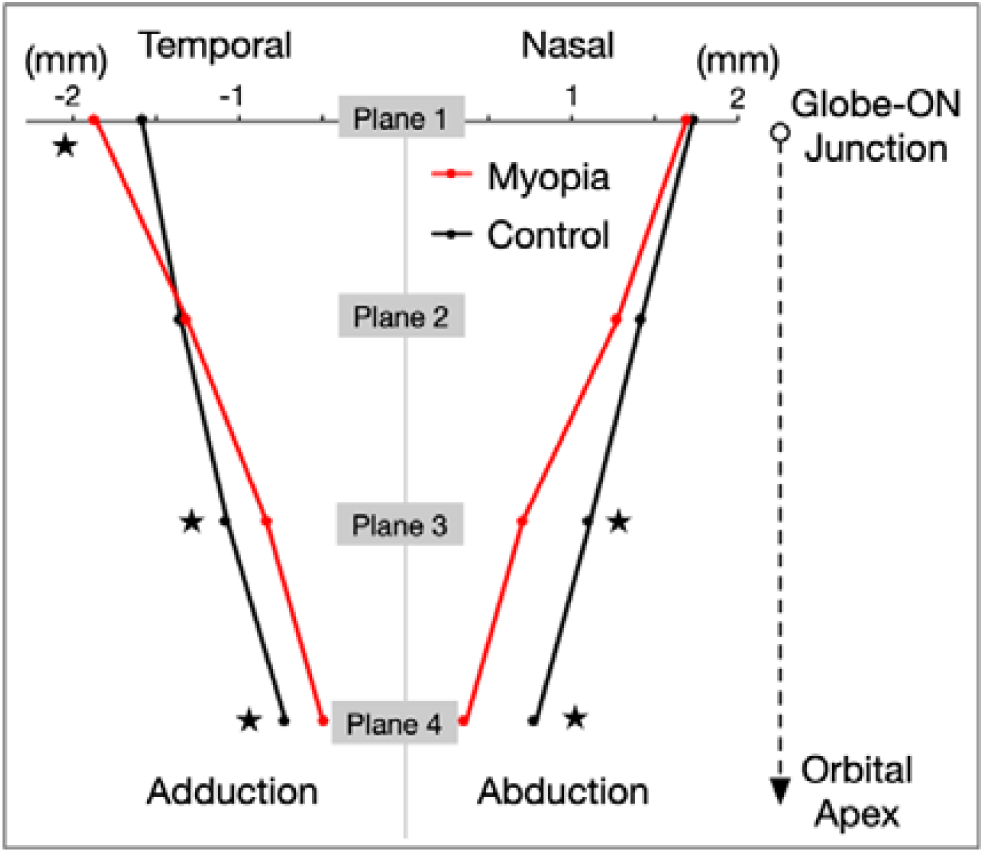
Optic nerve displacements in 4 quasicoronal planes in naso-temporal and supero-inferior directions. Optic nerve displacements were concentrated in the anterior segments for highly myopic eyes. ON: optic nerve.

### ON Insertion Angle

The ON insertion angle was 15.26 ± 3.72° for emmetropic and 25.68 ± 8.01° for HM eyes (p<0.001, **Figure 7A**). In myopic eyes, the ON insertion angle in Type-2 staphyloma was significantly different from eyes with no significant staphyloma (**Figure 7B**). The ON insertion angle was positively correlated with both ON tortuosity (**Figure 7C**) and with axial length (**Figure 7D**).

**Figure 7.**
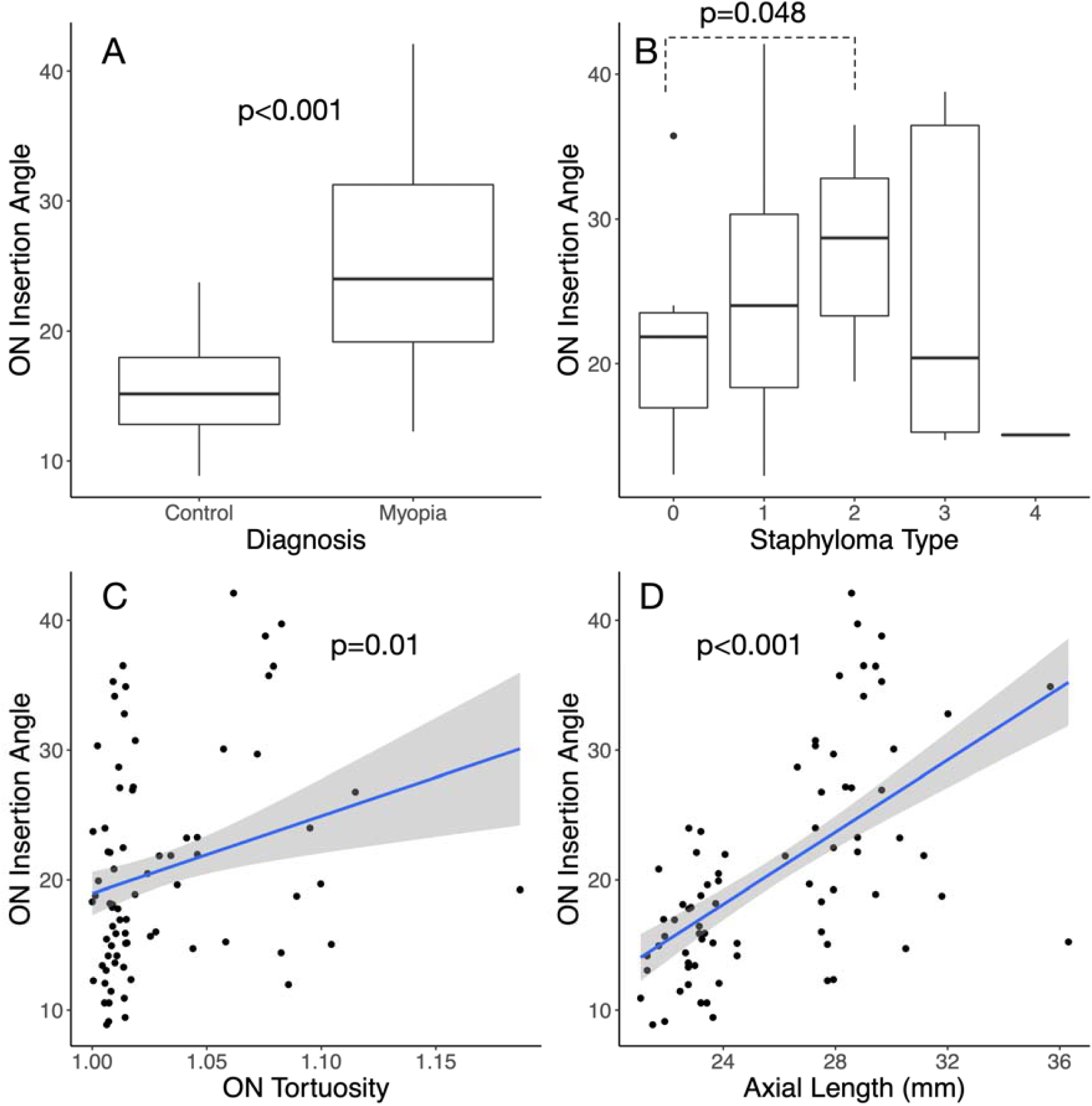
(**A**) Optic nerve (ON) insertion angle (in degrees) in control and high myopia groups. (**B**) ON insertion angle of high myopia eyes stratified by different staphyloma types. ON insertion angle was positively correlated with (**C**) ON tortuosity and (**D**) axial length.

In adduction, temporal translation of the globe increased with an increasing insertion angle (p = 0.006). Globe displacements in other directions were not correlated with ON insertion angle.

A larger insertion angle was correlated with a smaller ON displacements in plane 2 (adduction: p=0.04; abduction: p=0.009), plane 3 (adduction: p=0.03; abduction: p < 0.0001) and plane 4 (only in abduction: p=0.001).

## DISCUSSION

In this study, we evaluated the morphology and displacements of the globe and ON following horizontal eye movements in HM eyes and in healthy emmetropic controls. We found that the adjusted tortuosity of the ON was significantly lower in HM eyes, suggesting that eyes destined toward HM exhibited higher ON traction forces during eye movements before the onset of myopia. We have previously reported that the ON traction force was of the same order of magnitude as that of extraocular muscle forces and postulated that it may contribute to axial elongation in myopia.^21^ While we are unable to provide proofs to such speculations, our work provide a better understanding of the biomechanical forces acting on the posterior pole during eye movements in healthy and HM eyes.

In this study, we found that the adjusted ON tortuosity (i.e. the estimated tortuosity of the ON prior to the onset of myopia) was less in HM as compared to emmetropic control eyes, which may increase the ON traction force during eye movements if other factors remain unchanged. This could imply that at a prior point in time, a greater level of ON traction may have acted on the posterior pole thus encouraging axial elongation and myopia progression. However, at the present time, in HM eyes, the globe is excessively elongated, which therefore “relieved” some of the inherent ON traction. Even if at the present time HM eyes have more tortuous ONs than normal eyes, they may still be able to exhibit larger ON traction forces. For instance, for the same magnitude of eye movements, theoretically, the ON-globe junction needs to travel a longer distance in HM eyes when compared to normal eyes. This is indeed true in adduction from our measurements of ON location in the quasicoronal plane 1. Because the nonlinearity of ON traction force and orbital tissue resistance force, this protection effects of a more tortuous ON might be neutralized by a larger travel distance of ON-globe junction in HM eyes. Second, in HM eyes with staphyloma, the insertion of the ON onto the posterior scleral tends to be oblique (with the presence of a “tilted disc”). The location of the ON-globe junction would therefore be nasal to the posterior apex of the eye in most cases. This may cause the ON to wrap around the fulcrum of the posterior apex, especially in abduction for a large magnitude of eye movements. Additionally, depending on the location of the staphyloma, this may result in an even more extreme, posteriorly-displaced apex around which the ON must traverse during horizontal eye movements.

In this study, we chose to focus on the metric of ON disc tilt severity (objectively measured by assessing ON insertion angle), since it should reflect both axial length and distortion from staphyloma. We found that in HM eyes, the ON insertion angle in Type-2 (posterior pole) staphyloma was significantly greater than in HM eyes with no significant staphyloma (**Figure 7B**). This is consistent with the fact that the Type-1 (macular) staphylomas may have their ridges all temporal to the ON. Type-4 (peripapillary) and Type-3 (compound) staphyloma encompasses a great variety of possible ridge locations, so both of these types were limited by sample size for this particular analysis. We also found that the ON insertion angle was positively correlated with ON tortuosity (**Figure 7C**), axial length (**Figure 7D**) and temporal translation of the globe in adduction. Additionally, the ON insertion angle was negatively correlated with ON displacements in plane 2, plane 3 and plane 4 (only in abduction). These observations showed that the greater the disc tilt (and greater the insertion angle), the less taut the ON and the less the ON would move, suggesting that ON traction might be relieved by the increased in insertion angle. Therefore, it is possible that disc tilt is a compensatory mechanism of the eye to mitigate the effects of a large ON traction force during eye movements. It is also plausible that disc tilt is the consequence of continuous stretching of peripapillary tissues following repeated eye movements. The complexity of eye movement patterns in association with variations in eye and orbit anatomy may contribute to the variety of tilting directions and tilting severity among HM subjects, but this remains to be confirmed.

In emmetropic controls, the course of the optic nerve is oblique in the sagittal plane (**Figure 3**). Specifically, from the ON-globe junction, the ON travels towards the orbit apex posteriorly and superiorly, showing a typical ON course in the orbit. However, ONs were located more inferiorly in HM subjects compared with those in normal controls. It could be possible that in HM eyes, ONs were pushed posteriorly due to eye elongation and pulled inferiorly due to gravity (on average people spend 2/3 of their time with upright position). Under the combined effects of these two forces, the redundant ON moved inferiorly. The effects of ON location on the biomechanical environment of the globe in HM subjects are unknowns, and future computational modelling studies incorporating ON position may provide insights.

In emmetropic control eyes, the globe translated medially in adduction and laterally in abduction, which is in consistent with previous studies.^23^ However, in HM eyes, naso-temporal globe translation was significantly smaller than that in controls. It is possible that longer HM eyes experienced a larger resistance force from the orbital tissues during eye movements, resulting in a smaller globe translation. In the anterior-posterior direction, differences in globe translation between the two groups were not statistically significant although on average HM eyes had a larger globe retraction.

In adduction, ON displacements of HM subjects in planes 3 and 4 were smaller than those of emmetropic control eyes but the displacement of the ON-globe junction was larger, indicating that the displacements of the ON were more concentrated in the ON anterior segment in HM subjects. In abduction, a similar trend was observed. This should not be surprising, as the ON is relatively more “slack” in HM subjects. The pulling of the ON during eye movements would first eliminate the slack of the ON anterior segment before involving its most posterior part.

The ON, curved in 3D, exhibits a curvature in the sagittal plane and such a curvature differs between HM and emmetropic controls. From a pure mechanical point of view, in adduction or abduction, ON straightening in 3D could lead to vertical displacements of the ON. However, we found that the vertical displacements were almost null and no differences were found between HM and controls (**Table 1**). This indicates that horizontal eye movements only induce ON displacements in the horizontal plane. Therefore, a 2D axial slice of the eye should be sufficient to capture the ON displacements during horizontal eye movements. This is of high interest, because 2D MRI (rather than 3D) allows a much higher temporal resolution, enabling real-time imaging of eye movements.

As myopia is associated with near work, it has been proposed that accommodation is involved in myopia progression. Studies have also shown that convergence or other types of eye movement alone could induce transient axial elongation.^14–16^ Furthermore, reduction of convergence at near work by wearing prismatic lens is protective against myopic progression.^26^ Axial elongation during accommodation is likely related to the inward mechanical pulling force acting on the corneoscleral shell by the ciliary muscle. However, the mechanisms of convergence-induced axial elongation are unclear. We speculated that convergence-induced eye movements of both eyes could lead to a large ON traction force, especially in viewing near subjects.

In this study, several limitations warrant further discussion. First, age was not matched between two groups. It is possible that the degeneration of ocular tissues may affect ON displacement patterns. In spite of this limitation, we still believe this dataset provides us with an improved understanding of the behavior of ON displacements induced by eye movements. Second, we only evaluated the tortuosity and displacement of the anterior segment (15 mm long) of the intraorbital ON because of the low visibility of the posterior segment with MRI. Moreover, the full ON length was not measured. It is noted that the intraorbital ON length may be a very important factor that influences ON stretching during eye movement. Future studies should include this important parameter whenever possible. Third, eye rotation angles were 15° in this study. However, in the horizontal plane, human eyes can rotate up to ±50 ° in daily activities.^27^ For a larger magnitude of eye movements, the ON will be fully straightened and stretched. The effects of a larger magnitudes of eye movement need to be further explored.

In conclusion, the adjusted tortuosity of the ON was significantly lower in HM eyes, suggesting that eyes destined toward HM exhibited higher ON traction forces during eye movements before the onset of myopia. Our ON metrics may be valuable to explore a potential link between eye movements and axial elongation.

## Data Availability

Data available from the authors upon reasonable request.

## Supplementary material A

**Figure.**
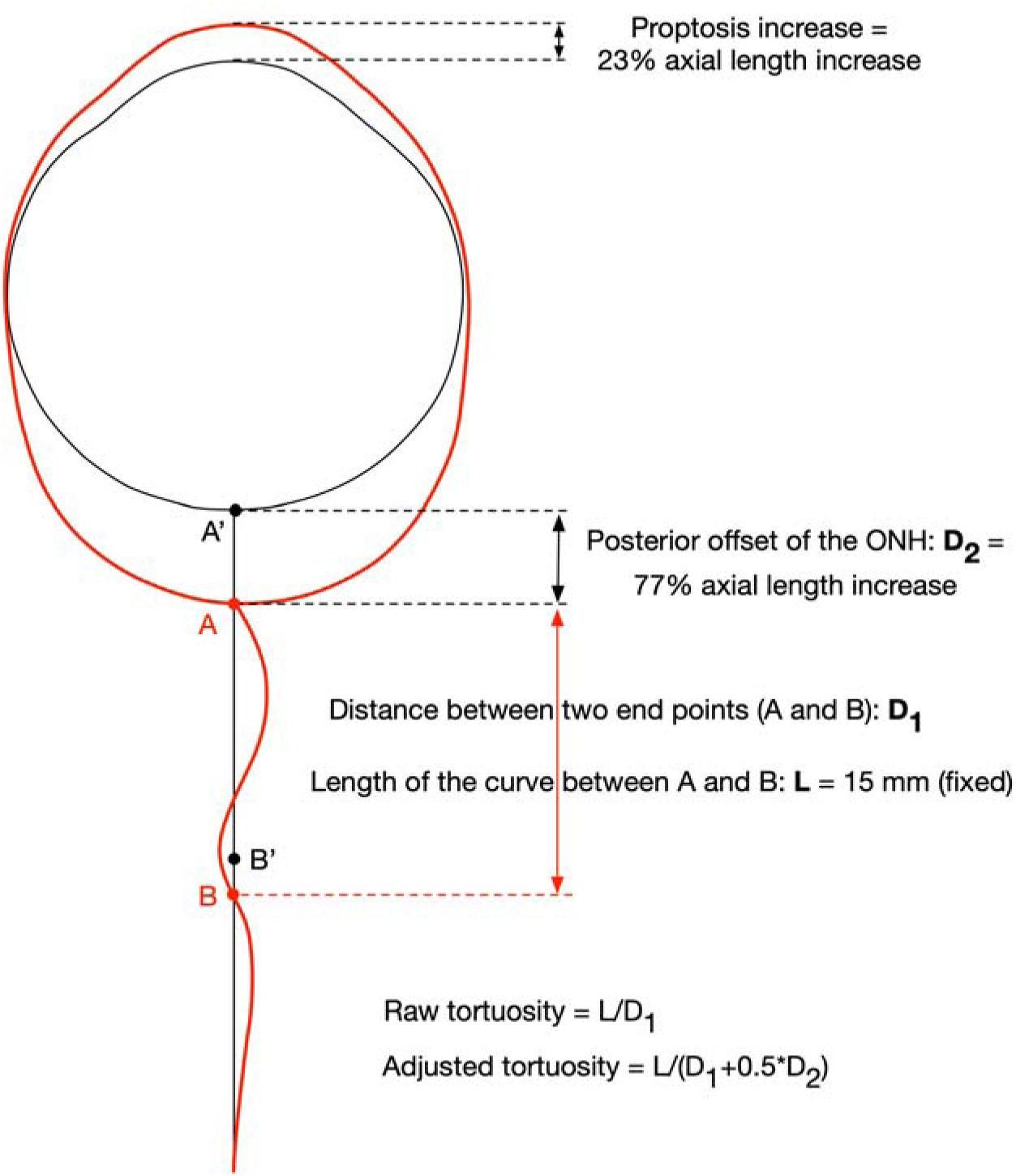
Illustration of ON tortuosity definitions. The red lines delineate a HM eye and the black lines represent the estimated configuration of the same eye prior to the onset of myopia at a prior point in time.

The tortuosity of the ON (non-adjusted) was defined as the length of the ON central line segment (15 mm long from the globe-ON junction) divided by the distance between the two end-points A and B (L/D_1_ in **Figure**).

The adjusted ON tortuosity was defined as the length of the ON central line segment (15 mm) divided by the distance between points A’ and B’. Points A’ and B’ were the hypothetical locations of points A and B at a prior point in time before the onset of myopia. The distance between A and A’ (D_2_) was set as 77% of the axial length increase compared to a normal eye (axial length: 22.84 mm) because proptosis also occurs during eye elongation.^1^ As only half of the whole ON was used in tortuosity calculations, the distance between B and B’ was set as half of the ONH displacement (D_2_) by assuming that the ON “stretch” is evenly distributed along the whole ON length (30 mm) when moving point A to A’.

Note that the tortuosity of the anterior segment of the ON is usually larger than that of the posterior half, thus the distance between B and B’ should be less than half of D_2_ or even near zero. Therefore, for a given HM eye, the adjusted tortuosity value is very conservative and should represent the upper limit of all possible tortuosity values prior to the onset of myopia.

